# Diversity in Parkinson’s disease genetics research: current landscape and future directions

**DOI:** 10.1101/2021.12.07.21266995

**Authors:** Artur F. Schumacher-Schuh, Andrei Bieger, Olaitan Okunoye, Kin Mok, Shen-Yang Lim, Soraya Bardien, Azlina Ahmad Annuar, Bruno Lopes-Santos, Matheus Zschornack Strelow, Mohamed Salama, Shilpa C Rao, Yared Zenebe Zewde, Saiesha Dindayal, Jihan Azar, LK Prashanth, Roopa Rajan, Alastair J Noyce, Njideka Okubadejo, Mie Rizig, Suzanne Lesage, Ignacio Mata, On behalf of the Global Parkinson’s Genetics Program (GP2)

**Author notes:** Corresponding author: Artur F. Schumacher-Schuh. Hospital de Clínicas de Porto Alegre, Serviço de Neurologia, sala 2040. Rua Ramiro Barcelos, 2350. CEP 90035-003. Porto Alegre, RS, Brazil. **Funding sources for study:** The Global Parkinson’s Genetics Program (GP2) is a resource project from the Aligning Science Across Parkinson’s (ASAP) initiative.

## Abstract

Human genetics research lacks diversity; over 80% of genome-wide association studies (GWAS) have been conducted on individuals of European ancestry. In addition to limiting insights regarding disease mechanisms, disproportionate representation can create disparities preventing equitable implementation of personalized medicine. This systematic review provides an overview of research involving Parkinson’s disease (PD) genetics in under-represented populations (URP), and sets a baseline to measure the future impact of current efforts in those populations.

We searched PubMed and EMBASE until October 2021 using search strings for “PD”, “genetics”, the main “URP”, and “lower-to-upper-middle-income countries”. Inclusion criteria were original studies, written in English, reporting genetic results on PD patients from non-European populations. Two levels of independent reviewers identified and extracted relevant information.

We observed considerable imbalances in PD genetic studies among URP. Asian participants from China were described in the majority of the articles published (61%), but other populations were less well studied, for example, Blacks were represented in just 4.0% of the publications. Also, although idiopathic PD was more studied than monogenic forms of the disease, most studies analyzed a limited number of genetic variants. We identified just seven studies using a genome-wide approach published up to 2021 including URP.

This review provides insight into the significant lack of population diversity in PD research highlighting the urgent need for better representation. The Global Parkinson’s Genetics Program (GP2) and similar initiatives aim to impact research in URP, and the early metrics presented here can be used to measure progress in the field of PD genetics in the future.

## Introduction

Since the Human Genome Project, the development of new technologies for the interrogation of genetic variability has increased exponentially, and new large-scale, high-throughput sequencing methods for genotyping and DNA sequencing have emerged, allowing thousands of genome-wide association studies (GWAS) to be performed. These technologies and the resulting analyses have revolutionized genetic investigation of disease, however, as pointed out by previous analyses of GWAS databases, these studies have failed in one major regard, they are not representative of the global genetic diversity. As a consequence of sample availability, budgetary constraints, issues with enrollment, or statistical power, populations of European ancestry still represent the majority of subjects included (Popejoy and Fullerton 2016; Hindorff et al. 2018). This lack of diversity has resulted in missed opportunities, such as the discovery of new genetic associations for complex traits and the discovery of novel genetic causes of monogenic forms of disease that could help to unveil unknown causes of these pathologies. It also threatens to jeopardize medical care, drug development, and advancements in precision medicine, preventing equitable health care among different populations (Hindorff, Bonham, and Ohno-Machado 2018; Sirugo, Williams, and Tishkoff 2019; Wojcik et al. 2019).

Parkinson’s disease (PD) is a multifactorial disorder in which a complex interaction between genetics and environmental factors occurs. As no curative or preventive therapy is currently available, exploring its pathophysiology is crucial to improve treatment. To date, approximately 20 genes with highly penetrant rare variants are linked to familial or monogenic forms of PD, predominantly among persons of European ancestry (Blauwendraat, Nalls, and Singleton 2020). A recent GWAS meta-analysis nominated 90 risk variants explaining approximately a quarter of the disease heritability. However, this study included just individuals of European ancestral origin, limiting the generalizability of these discoveries to other populations (Nalls et al. 2019). The largest PD GWAS among non-Europeans was recently reported in East Asians (Foo et al. 2020). The report included almost 7,000 individuals with PD and identified two novel risk loci. Research on PD genetics has increased in the last two decades, but a lack of diversity remains a significant problem for understanding the biological basis of the disease in all populations.

Many researchers are aware of the problem elicited by the lack of inclusion of under-represented populations (URP) and the hazards that result from avoiding or not achieving diversity within PD genetic studies. Nevertheless, most of the publications that raised this matter comprise comments, editorials, and letters, with only a few of them relying on empirical data (Khalil et al. 2020). Notwithstanding the value of these reports, which helped shed light on the issue, a deeper understanding of the geographic and ethnic coverage of PD genetic studies is necessary for building a solid roadmap for increasing diversity. This systematic review and bibliometric analysis aim to provide an overview of the publications in PD genetics in URP (individuals of non-European ancestry) to date, thereby clarifying the main gaps, identifying opportunities to ensure more diversity, and setting a baseline to measure the impact of future global efforts.

## Methods

We searched PubMed/MEDLINE and EMBASE from inception until October 2021. The search strings for each database were created using terms for “Parkinson’s disease”, “genetics”, “main non-European ethnic groups”, and the “lower to upper-middle-income countries”, as classified by the World Bank (Supplementary Table 1). Inclusion criteria were original studies reporting genetic results on PD from non-European populations and published in English. Systematic and narrative reviews, meta-analyses, and papers exclusively reporting functional, epigenetic, or biomarkers results were excluded.

The Rayyan software was used to detect duplicates and perform the first screening procedure (Ouzzani et al. 2016). We implemented the review in a two-step approach. First, two independent researchers screened titles and abstracts for inclusion criteria, and a third reviewer judged any discrepancies. Second, another reviewer examined the selected papers in their entire content to reassess inclusion criteria and collect data through an online extraction form. We collected information about the study design for each included study, classified as study of familial/monogenic cases, sporadic PD, or GWAS. Ethnicity was mainly categorized by geographical perspective, and we determined it based on the explicit description in the manuscript or inferred by the country of origin. Laboratory methods used for genetic analyses are very diverse, but to measure the access to technologies, we highlight those using next-generation sequencing (considered a “new” technology). The collaborative network was defined based on the number of distinct centers collecting samples and we classify them in single-center, multicenter within the same country or international multicenter. Finally, funding information was classified as funded exclusively from sponsors located in underrepresented regions or not.

Bibliometric analysis was also conducted, based on the titles previously selected that had a full record in the Web of Science Core Collection database. Based on this information, we retrieved the number of authors and citations per document, the impact factor of the journals, studies with authors from single or multiple countries, and the collaborative network among authors from different countries. For comparative analysis of the quality and visibility of the studies published, we reran the PubMed search without applying any exclusion criteria for the most productive countries in underrepresented regions. We compared the results with the same search for three different countries with mainly European ancestry from different continents (Germany, Canada, and Australia). Descriptive and comparative analyses between main ethnicities were performed in Python 3.9.5 and R 4.0.5, with the package ‘bibliometrix’ (Aria and Cuccurullo 2017).

## Results

After removing duplicates, we retrieved 1,606 titles from the search in PubMed/MEDLINE and EMBASE, from which 459 were excluded in the first screening step, resulting in 1,147 papers (Figure 1). In the second step, when the entire paper was examined, 180 papers were excluded, resulting in a final count of 967 (for a completed list of references included, see Supplementary Table 2). The main reasons for exclusion were studies not written in English (n = 74), 63 in Chinese, 7 in Spanish, 1 in Japanese, and 1 in Persian; none of them have performed a GWAS when examining their titles and abstracts. Other reasons for exclusions were not including individuals from URP, not including persons with PD, no genetic analysis performed and not reporting original results. For a subset of papers (n = 923) that were also available in the Web of Science Core Collection database, further bibliometric analysis was performed.

**Figure 1:**
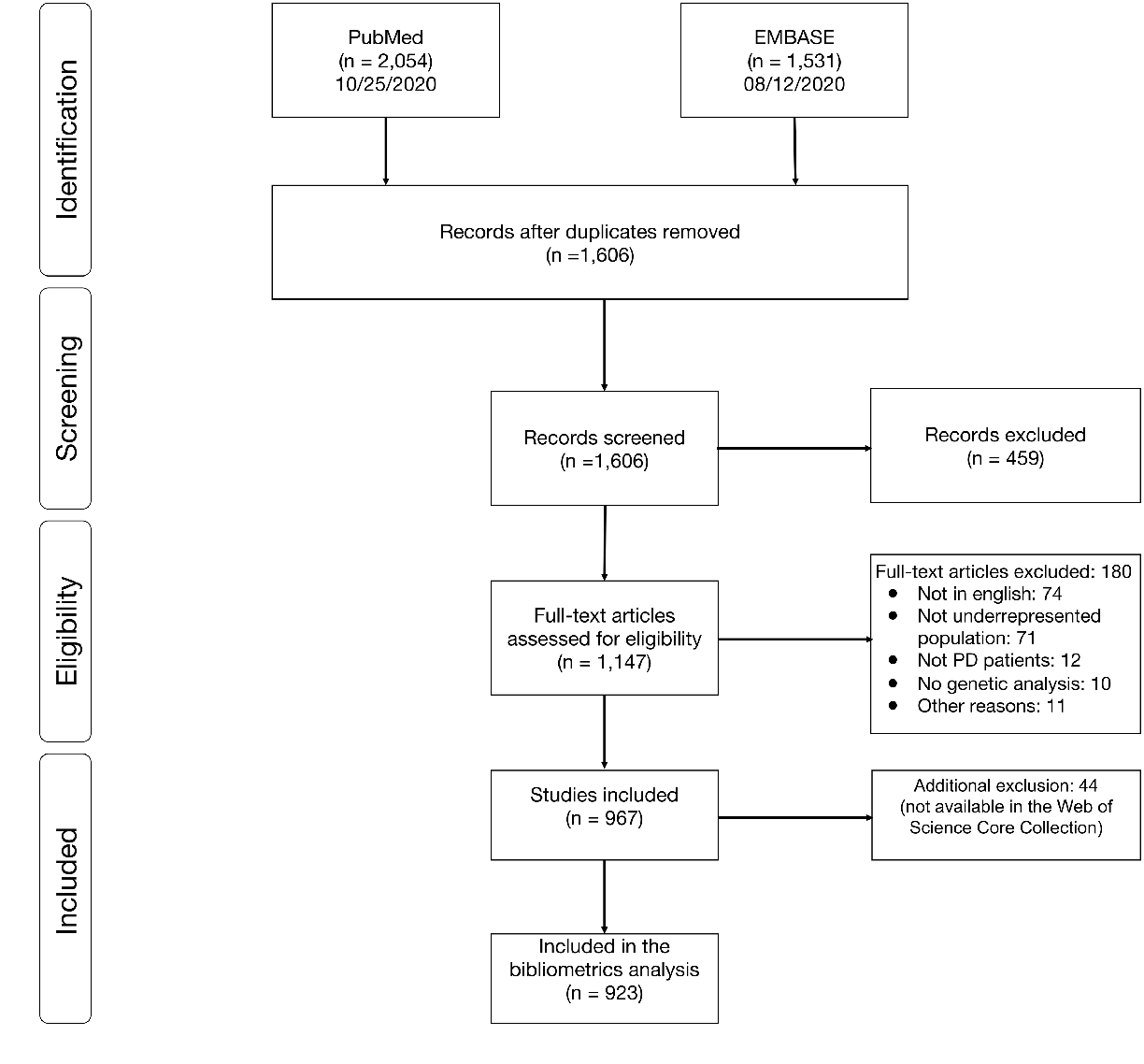
PRISMA flow diagram

The first two papers retrieved in our search were published in 1997, and we observed a trend of increasing publication counts each year, with 96 published in 2020 (Figure 2). However, the only consistent increase in publication counts along the years was among the Greater China region, while those among persons from Central and Southeast Asia, and Sub Saharan Africa or Black ancestry showed the lowest increases. Interestingly, we observe a decrease in publication counts for Latin America & Caribbean and the Middle East & North Africa in the previous five years. Overall, PD genetic publications from URP were dominated by participants from Chinese ancestry (n = 589, 60.9%), followed by participants from the Middle East & North Africa (n = 142, 14.7%), Latin America & Caribbean (n= 102, 10.6%), South Asia (n = 79, 8.2%), East Asia excluding Greater China (n = 69, 7.1%), and Sub Saharan Africa or other Blacks (n = 37, 3.9%). Just six publications were identified describing research from Central Asia.

**Figure 2:**
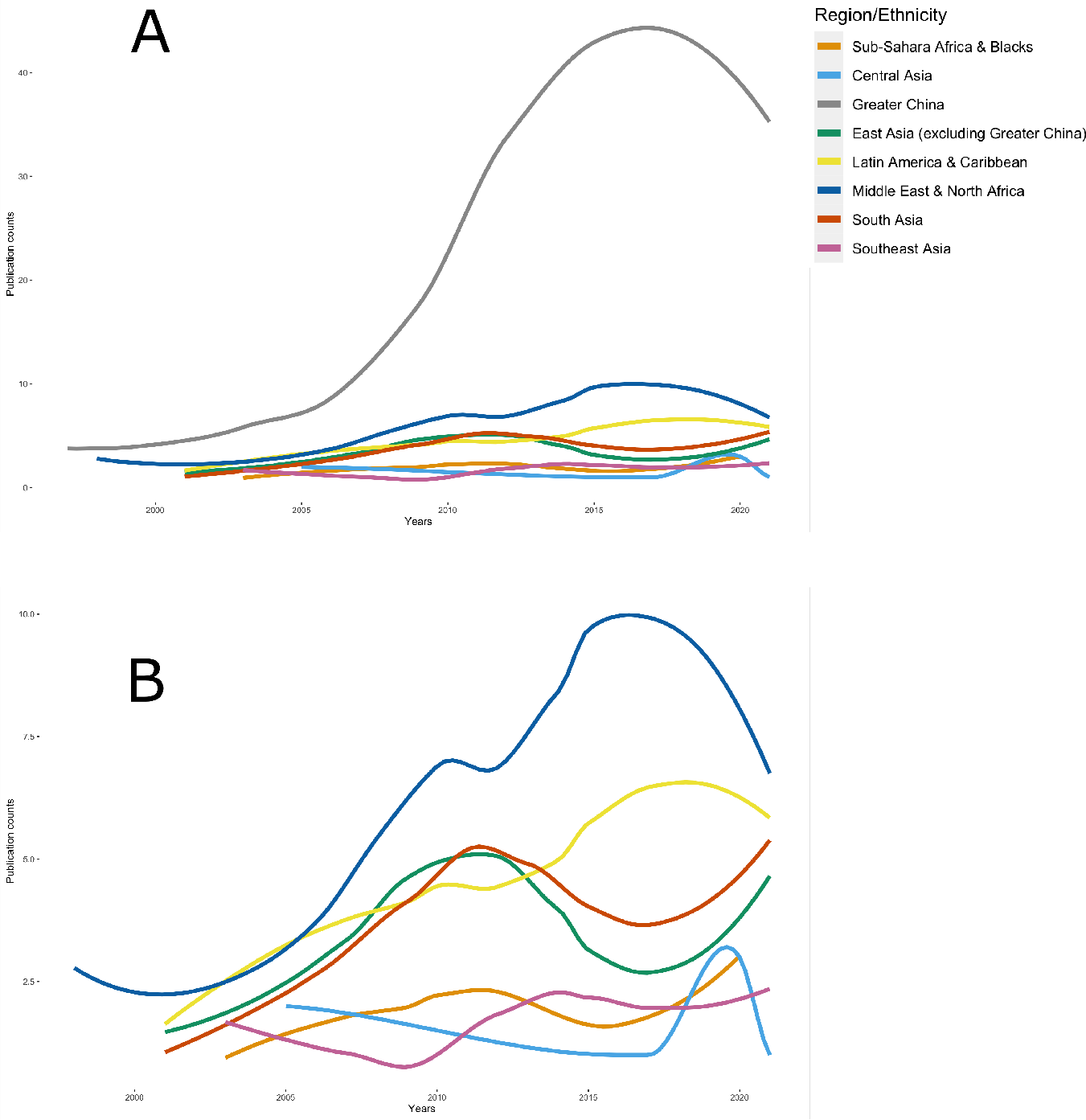
Publication counts along the years by each region/ethnicity A: All regions/ethnicity studied. B: Studies from Greater China were excluded to better visualize other regions/ethnicities. * Curves were smoothed by LOESS regression

Considering the corresponding authors’ countries/regions, which were not necessarily synonymous with the ethnic groups studied, the Greater China region has the highest number of papers (478, 51.8%), followed by the USA (60, 6.5%), India (54, 5.9%), Brazil (44, 4.8%), Japan (41, 4.5%) and Singapore (37, 4.0%) (Figure 3B). Studies including participants of Greater China ancestry were mainly from the Greater China region (477, 84.9%) and Singapore (34, 6.1%). East Asia excluding Greater China were mainly from Japan (40, 59.7%), and South Korea (12, 17.9%); South Asia from India (54, 70.1%) and South Africa (8, 10.4%); Southeast Asia from Singapore (11, 42.3%) and Malaysia (8, 30.1%); the Middle East & North Africa from USA (22, 16.8%), Iran (19, 14.5%), Israel (18, 13.7%) and France & Turkey (14, 10.7%, each); Latin America & Caribbean from Brazil (44, 43.6%), USA (19, 18.8%) and Mexico (16, 15.8%); Sub-Saharan Africa or other Blacks from South Africa (15, 41.7%), USA (9, 25%), France & Nigeria (3, 8.33%, each). All six papers from Central Asia were from correspondent author’s countries outside the region.

**Figure 3:**
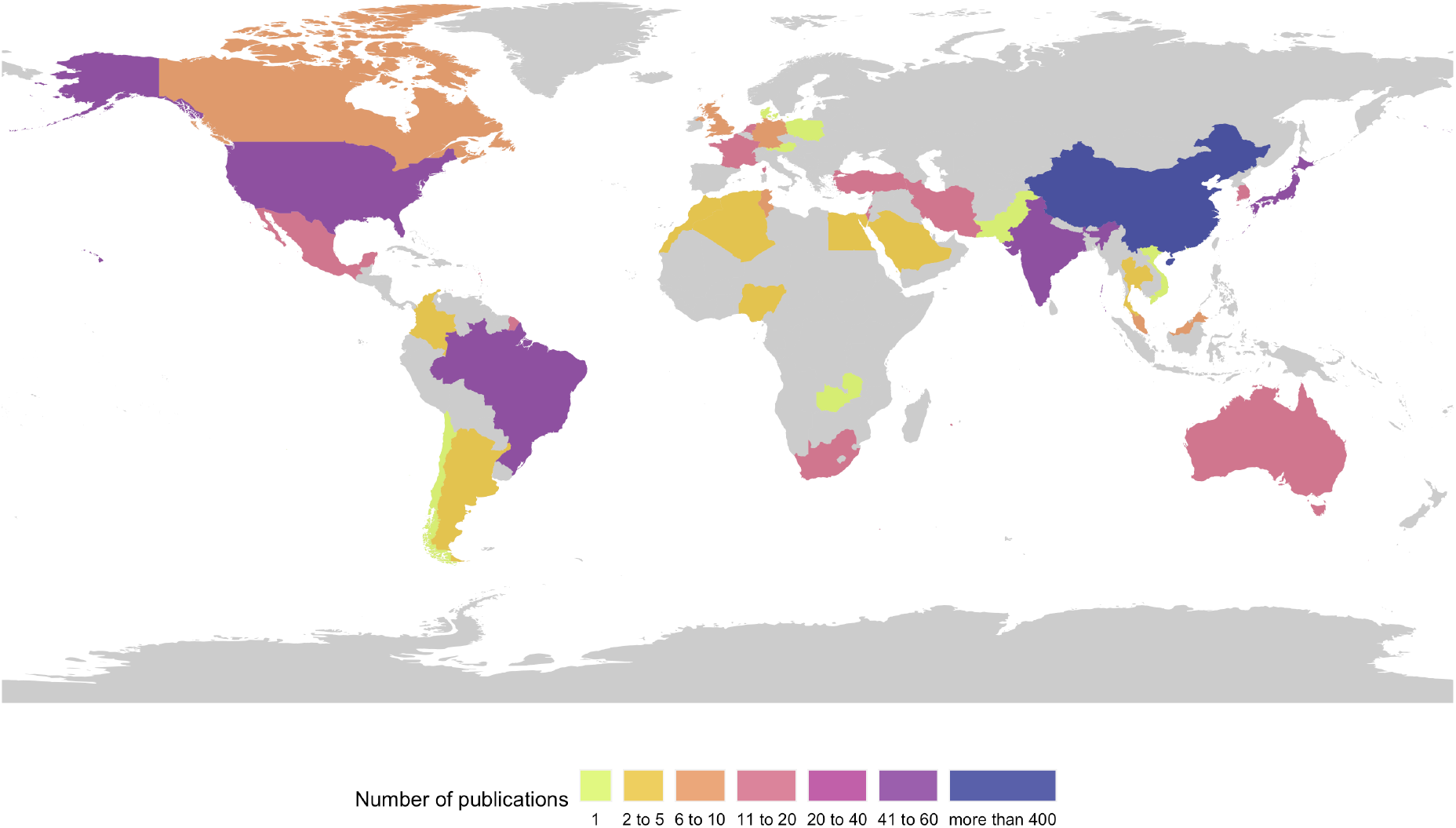
Publication counts by corresponding author’s countries/regions

Monogenic/familial forms of PD were the main focus of 211 (21.8%) studies, while the rest concentrated on case/control studies of genetic risk factors or genotype-phenotype associations of sporadic PD (Table 1). Greater China studies tended to focus on idiopathic PD, with less emphasis on monogenic/familial forms of PD (13.9%). In contrast, this latter type of study was more represented in the other groups, especially in South Asians (44.3%) and Sub-Saharan Africa or other Blacks (40.5%). GWAS were described only in 7 (0.7%) of the papers (Supplementary Table 3); one the most recent was published in 2020 and recruited almost 7,000 Asian patients for the discovery sample, being the largest cohort of PD patients among URP studies to date (Foo et al. 2020). Recently, a GWAS study among Latinos was published with almost 1,500 participants (Loesch et al. 2021). The use of newer technologies, such as next-generation sequencing, was performed in only 91 (9.4%) studies; mostly in Greater China ancestry (55, 9.3%). Latin America & Caribbean and Sub Saharan Africans & other Blacks presented the lowest relative use of this technology (4.9% and 5.4% respectively). Studies were funded exclusively by local resources in more than 60% of the studies in Asia (not including Central Asia), in Latin America, this number was approximately 52.9%, and in Africa, it dropped to 32.4%.

**Table 1:** Characteristics of the studies according to main ethnicities

|  | All studies | Chinese | East Asians (other than Chinese) | South Asians | Southeast Asians | Central Asians | Latin Americans & Caribbeans | Middle Easterns & North Africans | Sub Sahara Africans or other Blacks |
| --- | --- | --- | --- | --- | --- | --- | --- | --- | --- |
| Count* (%) | 967 | 589 (60.9) | 69 (7.1) | 79 (8.2) | 30 (3.1) | 6 (1) | 102 (10.6) | 142 (14.7) | 37 (3.9) |
| Average years from publication, mean (SD) | 7.3 (5.27) | 7.2 (5.35) | 8.45 (5.13) | 7.89 (5.2) | 7.87 (5.72) | 6.5 (7.56) | 8.29 (5.49) | 7.73 (5.22) | 9.0 (4.81) |
| Sample size, median (interquartile range) | 261.0 (106.0 - 515.75) | 384.0 (178.5 - 583.0) | 304.0 (171.0 - 741.0) | 154.0 (90.0 - 295.0) | 468.0 (156.25 - 690.0) | 107.0 (50.0 - 136.0) | 133.5 (79.5 - 219.25) | 132.0 (52.0 - 432.5) | 92.0 (39.0 - 210.0) |
| Authors per document, mean (SD) | 9.93 (7.53) | 9.21 (6.79) | 16.32 (12.12) | 7.09 (3.45) | 14.04 (11.34) | 18.33 (14.24) | 13.55 (13.03) | 13.27 (9.47) | 9.47 (8.85) |
| Citations per year per document, mean (SD) | 2.24 (4) | 2 (4.32) | 4.39 (11.18) | 1.63 (1.5) | 5.37 (16.89) | 6.11 (11.93) | 3.81 (9.33) | 4.25 (8.68) | 1.95 (1.36) |
| Impact factor, mean (SD) | 4.21 (4.75) | 3.86 (3.89) | 6.77 (9.73) | 3.45 (1.98) | 7.29 (14.27) | 5.35 (4.01) | 5.41 (9.8) | 6.55 (9.28) | 4.03 (2.21) |
| All authors from the same country, count (%) | 655 (71.04) | 428 (76.16) | 42 (62.69) | 65 (84.42) | 15 (57.69) | 1 (16.67) | 65 (64.36) | 47 (35.88) | 19 (52.78) |
| Studies with authors from multiple countries, count (%) | 267 (28.96) | 134 (23.84) | 25 (37.31) | 12 (15.58) | 11 (42.31) | 5 (83.33) | 36 (35.64) | 84 (64.12) | 17 (47.22) |
| Studies on monogenic PD, count (%) | 211 (21.82) | 82 (13.92) | 23 (33.33) | 35 (44.3) | 7 (23.33) | 2 (33.33) | 35 (34.31) | 46 (32.39) | 15 (40.54) |
| Exclusively funded by an URP country, count (%) | 635 (65.67) | 452 (76.74) | 42 (60.87) | 57 (72.15) | 25 (83.33) | 1 (16.67) | 54 (52.94) | 39 (27.46) | 12 (32.43) |
| Use of next generation sequencing technologies, count (%) | 91 (9.41) | 55 (9.34) | 10 (14.49) | 8 (10.13) | 3 (10.0) | 2 (33.33) | 5 (4.9) | 14 (9.86) | 2 (5.41) |
| Single center study, count (%) | 493 (50.98) | 344 (58.4) | 11 (15.94) | 39 (49.37) | 7 (23.33) | 1 (16.67) | 42 (41.18) | 50 (35.21) | 15 (40.54) |
| Multicenter,<br>same country,<br>count (%) | 216<br>(22.34) | 117<br>(19.86) | 20 (28.99) | 31 (39.24) | 15 (50.0) | 1 (16.67) | 25 (24.51) | 17 (11.97) | 9 (24.32) |
| Multicenter,<br>international,<br>count (%) | 112<br>(11.58) | 48 (8.15) | 18 (26.09) | 3 (3.8) | 6 (20.0) | 3 (50.0) | 26 (25.49) | 44 (30.99) | 8 (21.62) |
| Not clearly<br>stated, count<br>(%) | 142<br>(14.68) | 77 (13.07) | 19 (27.54) | 6 (7.59) | 2 (6.67) | 1 (16.67) | 9 (8.82) | 30 (21.13) | 5 (13.51) |

\*The same study may have included more than one ethnicity

For purposes of data collection, the largest number of the studies were conducted in a single-center (473, 51.8%), 216 (22.3%) had multiple centers within the same country, and 112 (11.6%) had multiple international centers. Single-center studies predominated in studies on participants of Greater China ancestry (58.4%), which presented a lesser proportion of international multicenter collaboration (8.2%). International multicenter studies were the lowest in South Asia (3.8%). A larger proportion of this type of study was observed in other regions, especially in the Middle East & North Africa (31%). Regarding the country of each co-author, most studies included authors from the same country (71.0%). Studies with authors from multiple countries were lowest among South Asia (15.6%) and Greater China (23.8%), and highest in Sub Saharan Africa & other Blacks (47.2%), the Middle East & North Africa (64.1%), and Central Asia (83.3%). In accordance with these results, the mean number of authors per publication was lowest in Greater China (9.2±6.8) and South Asia (7.1±3.5), higher in East Asia non-China (16.3±12.1), and highest in Central Asia (18.3±14.2). When examining the collaboration network (Figure 4), we observe that Sub-Saharan African and Asian countries intensely collaborated with Europe, while Latin American & Caribbean collaborated equally with North America. Collaborations between countries with a high proportion of underrepresented groups were limited, mainly within the same region.

**Figure 4:**
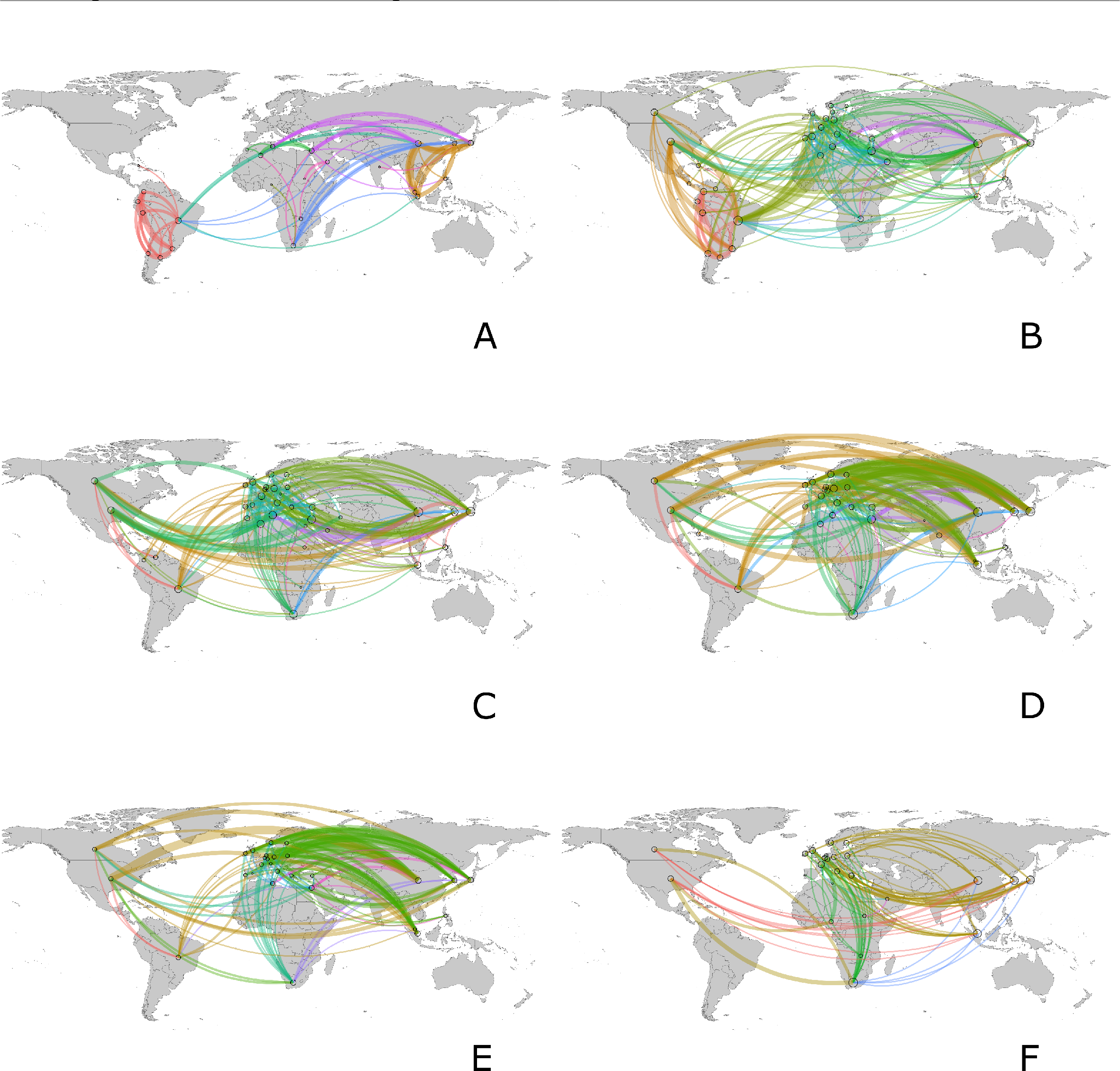
Collaboration among countries A: Collaboration map excluding the Global North, B: Collaboration map for articles describing Latin America & Caribbean populations, C: Collaboration map for articles describing Middle East & North Africa populations, D: Collaboration map for articles describing Asia excluding Greater China populations, E: Collaboration map for articles describing Greater China populations, F: Collaboration map for articles describing Subsahara Africa & other Black populations.

Southeast Asia presented the highest sample size (median 468), followed by China (median 384). Latin America & Caribbean and Sub Saharan Africa & other Blacks had the lowest sample size (medians 133 and 92, respectively). The highest citations per document were those from Central Asia (6.1±11.9), Southeast Asia (5.4±16.7), while East Asia excluding Greater China (4.4±11.1) and the Middle East & North Africa (4.3±8.7) presented intermediate citations. The lowest citation count was among Greater China (2±4.32), South Asia (1.63±1.5), and Sub-Saharan Africa & other Blacks (2.0±1.4). The average impact factor of the journals where the studies were published were highest in Southeast Asia (7.3±14.3), East Asia excluding Greater China (6.8±9.7) and the Middle East & North Africa (6.6±9.3) and lowest in Greater China (3.9±3.9) and South Asia (3.5±2.0). In comparison to the three countries with a predominance of European ancestry, countries with a predominance of URP publish in journals with a lower impact factor and obtain fewer citations (Figure 5).

**Figure 5:**
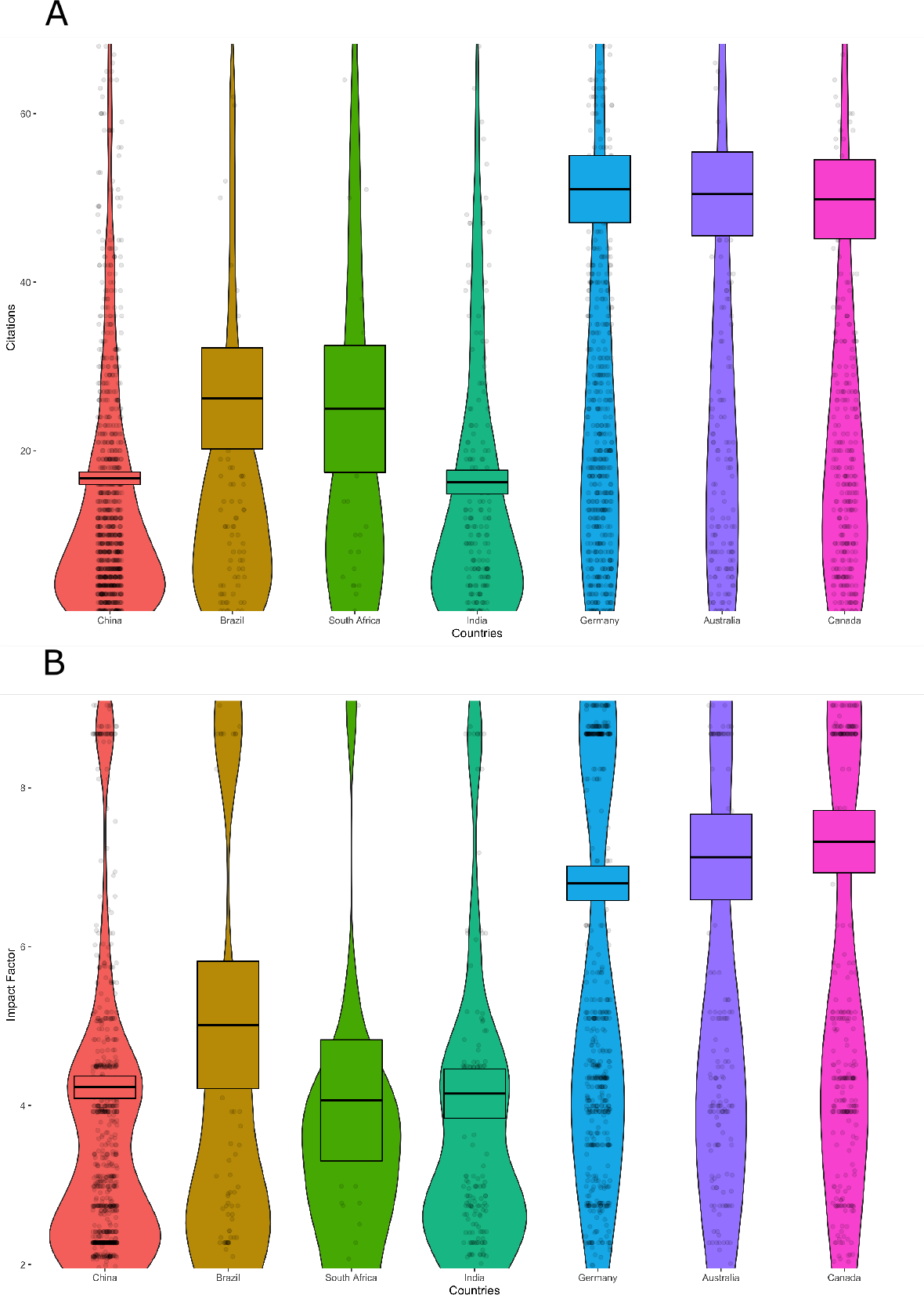
Citations and journal’s impact factor across selected countries A: Number of citations by country; B: Journal’s impact factor by country

## Discussion

This review aimed to provide an overview of the current situation for PD genetics research among URP, identify strengths and limitations, outline critical directions for future efforts and set a baseline to measure their impact. We believe that the summary provided here represents a significant step forward to highlight disparities and foster representativeness, with the potential to prevent inequalities in the healthcare of PD patients. Notably, we observed considerable imbalances in PD genetic studies among URP. While Greater China was described in the majority of the articles published (61.6%), other ethnic groups were less well studied, for example, Sub-Saharan Africans & other Blacks representing just 4% of the publications. Although idiopathic PD was more studied than monogenic forms of the disease, most studies analyzed a limited number of genetic variants. We identified just seven studies using a genome-wide approach published until 2021.

The bias towards European ancestry populations is a well-established problem in genetics, especially in GWAS. Efforts have been initiated to address this by major research funders, but we are still far from the desired equity (Siddiqi and Koemeter-Cox 2021). To further understand this bias, our search focused on PD genetic studies performed in non-European populations, which account for a great variety of ethnic backgrounds. Chinese populations are represented in 61.6% of the studies, with the Greater China region having half of the papers published in the field (452, 49.5%), followed by the Middle East & North Africa and Latin America & Caribbean. In addition, we observed that publication counts in Greater China are increasing on an annual basis, which is consistent with Popejoy et al. (2016), who stated that these groups were the most effective in their efforts to improve representation in genetic research (Popejoy and Fullerton 2016). Besides having a vast population compared to the rest of the world, their economic growth with significant investments in science and education can also explain this progress. For example, China increased more than 10% of its expenditure on science in 2020 (Jia 2020). On the other hand, Central Asia had just four papers published, all of them from researchers based in foreign countries. Black ancestry was represented in just 4% of the studies, without a perceptible trend of increase in the publication counts over the years. The majority of Blacks reside in sub-Saharan Africa and Latin America, both regions with countries within the lower-income and lower-middle-income strata. Consequently, economic constraints foster the limited expenditure on and development of research and innovation in general, and specifically for non-communicable diseases such as PD. Another barrier in many parts of the region is the access to specialized healthcare, such as neurologists, to ascertain disease diagnosis. Finally, historical discrimination and misconceptions about the purpose of research also contribute to lower participation rates of this group in research studies.

The highest frequency of studies on idiopathic PD was in Greater China, with all the other populations showing a higher frequency of studies on monogenic forms of PD. They also presented the highest median sample size and a higher proportion of the studies funded exclusively by local resources. Studying a multifactorial disorder such as idiopathic PD is a logistical and financial challenge since large samples need to be recruited to have sufficient power to detect minor effects and control for confounders. Probably, studying monogenic forms is often a more straightforward endeavor for lower-income countries since fewer participants are needed to be recruited. The genetic analysis for such rare forms, although costly, can also usually be done in partnership with laboratories from higher-income countries. Following this observation and considering that the sample size can be an indirect indicator of study quality, we see that studies with Asian populations, especially Greater China, reported the largest sample size. However, regarding citations per publication and the journal’s impact factor, another indirect index of quality, Greater China presented lower figures than other regions. Studies in the Greater China population have mainly investigated candidate genes in idiopathic PD, and to date, more comprehensive study designs like GWAS studies that can potentially generate more citations are still infrequent. Another potential explanation for this lower citation rate is that Greater China scientific publications have generally been more recent compared to the others. Southeast Asia is an exceptional case. Despite its still lower number of publications, they were able to recruit the largest sample sizes in individual studies, most of them locally funded, and exhibiting the highest citations and impact factor. Countries with a high economic development are in this region, such as Singapore and Malaysia.

Collaborative studies are crucial in genetics when we need to gather a substantial sample size. Besides that, a research network can be beneficial for underserved countries since it can strengthen credibility, facilitate data sharing, and promote capacity building. Most of the samples collected for PD genetic studies in South Asia and Greater China populations were from single centers, and international collaborations were scarce. In South and Southeast Asia, there were more local multicenter studies, but still limited international collaborations. These observations are supported by other collaborative indicators, such as the number of authors per publication and the frequency of studies with authors from a single country, both lower among Asians. This trend might be partially explained by a higher research capacity in Asia, especially in East and Southeast Asia. Stringent local regulations that govern data and biospecimen sharing, while intended to prioritize and develop local capacities, may also limit extensive international collaborations in several regions. Other factors that may further contribute to the limited collaborations might include language barriers, and cultural issues discouraging collaborations. The highest indicators of collaboration were observed in the Middle East & North Africa studies, with more international multicenter studies and increased frequency of authors from multiple countries. A possible explanation for this is the higher frequency of *LRRK2* p.G2019S carriers in this region, which could have piqued international interest (due to its common occurrence in European and Ashkenazi Jewish populations) and fostered international collaborations (Tolosa et al. 2020; Lesage et al. 2009). The need for high investment capital in cutting-edge technologies might also promote collaborations with higher-income countries.

A notable limitation of the present study can be the number of publications used as the primary measure of population representation. One could also argue that the broad search criteria used include such a diversity of studies that their joint numbers express vague concepts. Even so, we consider that our approach provides key indirect measures of scientific interest, development, and output in those specific populations, which reflect representation not only at the DNA level in databases but also at a much broader appraisal of the population’s social, financial, and scientific aspects. It is also important to note that many studies relied on the same samples for their analyses, thus not adding diversity at the DNA level, and increasing the chance of error by performing multiple comparisons. Furthermore, a thorough quality assessment of each study was not performed since we did not find an objective assessment tool, such as the CONSORT statement for clinical trials, which covers all the different study types included in our search. Also, this task would require extensive effort, which could prevent its replication in the future. Instead, we used indirect measures to estimate quality and also visibility, like citations and the journal’s impact factor. Finally, another limitation was the inclusion of only English-written publications. Although possibly introducing selection bias for higher impact papers, this choice narrowed the analysis to articles with higher international visibility.

As pointed out by Popejoy & Fullerton (2016), we believe future efforts to increase diversity in genomic research should include both bottom-up and top-down strategies. Researchers should acknowledge the importance of diversity in their studies, formulating questions and proposing robust study designs considering genomic diversity and its relationship with socioeconomic and environmental factors. Strategies to ensure recruitment among populations not used to participating in research include engaging local communities and proposing solutions to improve health care. From an analytical perspective, increasing information gain can be achieved through tools like trans-ethnic fine-mapping (Li and Keating 2014). On the other hand, funding agencies should promote representation by providing dedicated funding, increasing diversity among researchers, and applying knowledge to healthcare systems (Siddiqi and Koemeter-Cox 2021). Capacity-building in countries with a predominance of URP is another crucial step to guarantee long-term studies, and build autonomy and a diverse pool of researchers with expertise in genetics research (Lim et al. 2019; Hamid et al. 2021; Rizig et al. 2021). As observed in our results, there is a surplus of opportunities for promoting sustainable diversity in genomics studies through the empowerment of local researchers and authorities. This could be achieved either through collaboration with higher-income countries or by designing plans for regional development with specialized centers. International institutions have a key role in creating common fora for partnership development with higher-income countries or organizing underserved countries for regional ventures. Moreover, journals and editors should be sensitive about the importance of increasing diversity in publications, and a first step would be to increase representativeness in editorial boards. Editions focused on underrepresented populations, and specific criteria for publication could also be considered. Finally, peer-review should be carefully conducted to be a supportive and productive process fostering diversity.

The Global Parkinson’s Genetics Program (GP2, http://gp2.org/) is a project that aims to provide a comprehensive understanding of the genetic architecture of PD, utilizing strategies that include collecting large-scale data from URP worldwide and enabling researchers from those populations to drive this work forward (Global Parkinson’s Genetics Program 2021). To accomplish this ambitious goal, GP2 established a specific working group (the Underrepresented Populations Working Group; https://www.gp2.org/working-groups/underrepresented-populations-working-group/) comprising researchers from different countries and ethnic backgrounds to ensure adequate global representation. For data collection, GP2 is creating a consortium and projects to recruit subjects from URP inside the US called the Black and African American Connections to Parkinson’s Disease Study (BLAAC PD). Already existing initiatives in East Asia (IPDGC-Asia), India (LUX-Giant), Latin America (LARGE-PD), and Africa (IPDGC-Africa) receive strong support from the program. Besides data collection, strategies for collaborative data upload, access, and analysis are making it possible for GP2 to perform projects such as a trans-ethnic meta-analysis and fine-mapping, involving a diversity of researchers, resources, and data. To foster collaboration and build resources, GP2 offers a wide range of training opportunities for researchers around the globe, including online courses, Masters degrees and Ph.D. programs, and short-duration training visits on genetics and bioinformatics.

In conclusion, although steps have been taken worldwide to ensure diversity in PD genetic studies, the unbalanced efforts between URP are still concerning, as highlighted here. Among growing economies, we observed a steady increase in publications over the years, while this rate has been slower in lower-income regions. Concerted efforts are needed to recognize diversity as a driver of equality and scientific discoveries. Researchers, universities and funders, either public or private, should assume more active roles in paving the paths to achieve sustainable diversity through joint efforts, capacity-building, training, data sharing, and consciously redirecting capital. In this sense, the GP2 is playing an ambitious role in unveiling PD’s genetic architecture by engaging leaders, researchers, and study participants from Africa, Asia, Middle East, Latin America, and all other underrepresented populations within more developed nations. Hopefully, within the next few years, we will see a more inclusive research environment being translated into more and higher quality publications among under-represented groups, with potential parallel improvements in the healthcare of all populations.

## Data Availability

All data produced in the present study are available upon reasonable request to the authors

