## Supplementary table 1 for "Diversity in Parkinson’s disease genetics research: current landscape and future directions"

Supplementary Table 1: Detailed search string.

| Database | Search string |
|---|---|
| PubMed | ("parkinson disease"[MeSH Terms]) AND (gene OR genetic* OR genomic*) NOT (systematic[sb] OR Review[ptyp] OR Letter[ptyp] OR Editorial[ptyp]) AND Humans [filter]<br>AND<br>((latino OR hispanic* OR africa OR black OR asia*)<br>OR<br>(American Samoa OR China OR Fiji OR Micronesia OR Indonesia OR Cambodia OR Kiribati OR Laos OR Marshall Islands OR Myanmar OR Mongolia OR Malaysia OR Nauru OR Philippines OR Papua New Guinea OR North Korea OR Solomon Islands OR Thailand OR Timor-Leste OR Tonga OR Tuvalu OR Vietnam OR Vanuatu OR Samoa OR Djibouti OR Algeria OR Egypt OR Iran OR Iraq OR Jordan OR Lebanon OR Libya OR Morocco OR West Bank and Gaza OR Syria OR Tunisia OR Yemen OR Angola OR Burundi OR Benin OR Burkina Faso OR Botswana OR Central African Republic OR Cote d'Ivoire OR Cameroon OR Democratic Republic of the Congo OR Republic of the Congo OR Comoros OR Cabo Verde OR Eritrea OR Ethiopia OR Gabon OR Ghana OR Guinea OR Gambia OR Guinea-Bissau OR Equatorial Guinea OR Kenya OR Liberia OR Lesotho OR Madagascar OR Mali OR Mozambique OR Mauritania OR Mauritius OR Malawi OR Namibia OR Niger OR Nigeria OR Rwanda OR Sudan OR Senegal OR Sierra Leone OR Somalia OR South Sudan OR Sao Tome and Principe OR Eswatini OR Chad OR Togo OR Tanzania OR Uganda OR South Africa OR Zambia OR Zimbabwe OR Afghanistan OR Bangladesh OR Bhutan OR India OR Sri Lanka OR Maldives OR Nepal OR Pakistan OR Argentina OR Belize OR Bolivia OR Brazil OR Colombia OR Costa Rica OR Cuba OR Dominica OR Dominican Republic OR Ecuador OR Grenada OR Guatemala OR Guyana OR Honduras OR Haiti OR Jamaica OR Saint Lucia OR Mexico OR Nicaragua OR Peru OR Paraguay OR El Salvador OR Suriname OR Saint Vincent and the Grenadines OR Venezuela OR Albania OR Armenia OR Azerbaijan OR Bulgaria OR Bosnia and Herzegovina OR Belarus OR Georgia OR Kazakhstan OR Kyrgyz Republic OR Moldova OR North Macedonia OR Montenegro OR Romania OR Russian Federation OR Serbia OR Tajikistan OR Turkmenistan OR Turkey OR Ukraine OR Uzbekistan OR Kosovo)) |
| EMBASE | 'parkinson disease' AND ('latino'/exp OR latino OR hispanic* OR 'africa'/exp OR africa OR 'black'/exp OR black OR asia* OR 'american samoa' OR 'china' OR 'fiji' OR 'micronesia' OR 'indonesia' OR 'cambodia' OR 'kiribati' OR 'laos' OR 'marshall islands' OR 'myanmar' OR 'mongolia' OR 'malaysia' OR 'nauru' OR 'philippines' OR 'papua new guinea' OR 'nth kea' OR 'solomon islands' OR 'thailand' OR 'tim-leste' OR 'tonga' OR 'tuvalu' OR 'vietnam' OR 'vanuatu' OR 'samoa' OR 'djibouti' OR 'algeria' OR 'egypt' OR 'iran' OR 'iraq' OR 'jdan' OR 'lebanon' OR 'libya' OR 'mocco' OR 'west bank and gaza' OR 'syria' OR 'tunisia' OR 'yemen' OR 'angola' OR 'burundi' OR 'benin' OR 'burkina faso' OR 'botswana' OR 'central african republic' OR 'cote ivoire' OR 'cameroon' OR 'democratic republic of the congo' OR 'republic of the congo' OR 'comos' OR 'cabo verde' OR 'eritrea' OR 'ethiopia' OR 'gabon' OR 'ghana' OR 'guinea' OR |

| | |
|---|---|
| | 'gambia' OR 'guinea-bissau' OR 'equatial guinea' OR 'kenya' OR 'liberia' OR 'lesotho' OR 'madagascar' OR 'mali' OR 'mozambique' OR 'mauritania' OR 'mauritius' OR 'malawi' OR 'namibia' OR 'niger' OR 'nigeria' OR 'rwanda' OR 'sudan' OR 'senegal' OR 'sierra leone' OR 'somalia' OR 'south sudan' OR 'sao tome and principe' OR 'eswatini' OR 'chad' OR 'togo' OR 'tanzania' OR 'uganda' OR 'south africa' OR 'zambia' OR 'zimbabwe' OR 'afghanistan' OR 'bangladesh' OR 'bhutan' OR 'india' OR 'sri lanka' OR 'maldives' OR 'nepal' OR 'pakistan' OR 'argentina' OR 'belize' OR 'bolivia' OR 'brazil' OR 'colombia' OR 'costa rica' OR 'cuba' OR 'dominica' OR 'dominican republic' OR 'ecuad' OR 'grenada' OR 'guatemala' OR 'guyana' OR 'honduras' OR 'haiti' OR 'jamaica' OR 'saint lucia' OR 'mexico' OR 'nicaragua' OR 'peru' OR 'paraguay' OR 'el salvad' OR 'suriname' OR 'saint vincent and the grenadines' OR 'venezuela') AND (genetic OR gene OR genomic) AND [article]/lim AND [humans]/lim AND [clinical study]/lim |
